# Apixaban drug level monitoring in hemodialysis

**DOI:** 10.1101/2023.06.13.23291319

**Authors:** Simeon Schietzel, Andreas Limacher, Matthias B. Moor, Cecilia Czerlau, Bruno Vogt, Fabienne Aregger, Dominik E. Uehlinger

**Affiliations:** Division of Nephrology and Hypertension, Inselspital, Bern University Hospital, University of Bern, Bern, Switzerland; TU Bern, University of Bern, Bern, Switzerland; Division of Nephrology, Central Hospital Biel, Biel, Switzerland

**Keywords:** Apixaban, hemodialysis, drug, level, monitoring, anticoagulation

## Abstract

**Background:** Apixaban is increasingly being used in hemodialysis patients. However, uncertainty remains regarding appropriate dosing and risk of accumulation.

**Methods:** We analyzed apixaban drug levels from a tertiary care dialysis unit collected between August 2017 and January 2023. We compared 2.5 mg once versus twice daily dosing. We applied mixed-effects models analyses including dialysis modality, adjusted standard Kt/V, ultrafiltration and dialyzer characteristics.

**Results:** We analyzed 143 apixaban drug levels from 24 patients. Mean (SD) age was 64.2 (15.3) years (45.2% female), median (IQR) follow up 12.5 (5.5 - 21) months. For the 2.5 mg once and twice daily groups, median (IQR) drug levels were 54.4 (< 40 - 72.1) and 71.3 (48.8 - 104.1) ng/mL (P < 0.001). Drug levels were below the detection limit in 30 % and 14 %. Only dosing group (twice versus once daily) was independently associated with higher drug levels (P = 0.002). Follow-up did not suggest accumulation. 95^th^ percentile did not exceed those of non-CKD populations taking 5 mg twice daily. Drug levels before a bleeding (8 episodes) were significantly higher than those without a subsequent bleeding: 115 (SD 51.6) versus 65.9 (SD 31.6) ng/mL (P < 0.001). Patients with versus without a bleeding took concomitant antiplatelet therapy in 86% versus 6% (P < 0.001). In 21% of patients, drug level monitoring resulted in change of dosing.

**Conclusion:** Apixaban drug monitoring might be a contributory tool to increase safety in patients on hemodialysis. Further prospective outcome studies are warranted to investigate possible target levels.

## Introduction

Apixaban is increasingly being used in hemodialysis patients for prevention and therapy of thrombotic events. However, there is still uncertainty with respect to appropriate dosing and risk factors for accumulation.

Considerable non-renal excretion (73 %),^1^ lack of active metabolites,^2^ partial removal by hemodialysis (AUC -14 to -48 %) ^3, 4^ as well as favorable cohort and clinical trial data,^5^ make apixaban a valid alternative to vitamin K antagonists in hemodialysis. Guidelines of the American Heart Association, the American College of Cardiology and the Heart Rhythm Society (AHA/ACC/HRS) referring to apixaban as a reasonable direct oral anticoagulant alternative to warfarin in patients with atrial fibrillation who have end stage kidney disease (ESKD) or are on dialysis.^6^

However, plasma protein binding is high (87%)^7^ and the volume of distribution is large (21L/Kg)^1^ increasing the risk of accumulation in patient with reduced kidney function. The European Medicines Agency does not recommend apixaban in ESKD or dialysis^8^.

For patients on hemodialysis, the FDA recommends the standard dosing of 5 mg twice daily, with reduced dose (2.5 mg twice daily) only if an additional factor, older age (≥ 80 years) or lower weight (≤ 60 Kg) is present.^9^ FDA recommendations were based on a small pharmacokinetic study,^3^ reflecting the scarcity of data in the hemodialysis population.

A clinical trial with 97 patients on hemodialysis demonstrated equivalence to vitamin K antagonists at reduced doses (2.5 mg twice daily).^10^ However, with reduced dosing, a large retrospective cohort study found increased risks of systemic embolism and death compared to standard dosing or warfarin without increasing bleeding risks at standard dosing^11^. The AHA/ACC/HRS rather advocate for the reduced dose of 2.5 mg twice daily for patients on hemodialysis also emphasizing the lack of data.^6^ Even with reduced doses, serious bleeding events have been reported.^12^ As inter-individual variability of drug level is high,^1^ even 2.5 mg once daily dosing might be considered for individual prophylactic indications.

As a result, uncertainty remains regarding appropriate dosing strategies, which might be reflected by frequent off-label under-dosing.^13^

Drug monitoring is a legitimate approach to approximate a patient’s risk of an overtly strong or rather weak exposure to anticoagulant effect under a given dose.^8^ However, studies providing comparative guidance for different dosing regimens in patients on hemodialysis are scarce.

Age, sex, body weight and kidney function have been shown to impact apixaban drug half-life.^1^ It is unclear, if these factors apply to hemodialysis patients. No long-term investigations of apixaban level in hemodialysis patients are available.

We provide results from over 5 years apixaban drug monitoring. We investigate possible individual and dialysis-related risk factors for accumulation and provide comparative analyses to non-CKD populations. In addition, we include an exploratory analysis regarding drug levels and bleeding events.

## Methods

### Population

Chronic hemodialysis patients from a tertiary care center at the university hospital of Bern, Switzerland.

### Study type

Retrospective analysis of 5.5 years health-related data, available from routine clinical practice.

### In- and exclusion criteria

We included all adult patients under apixaban treatment at the initiation of the study that gave their informed consent to further use of health-related data. Inclusion was independent of apixaban dosing or indication. We excluded patients on peritoneal dialysis and those on home hemodialysis. Only scheduled apixaban drug level measurements were included. We excluded drug level that had been taken in other departments, during emergencies or before surgical procedures.

### Measurement of apixaban

The unit’s drug monitoring program was set to measure apixaban drug levels on a regular base targeting measurements every three to six months but allowing for individual adjustments of the measurement intervals according to the treating physician incorporating previous values and patient factors. Measurements were performed at the beginning of the week (long interval) before the start of the dialysis treatment.

Blood samples were obtained before administration of anticoagulants. After discarding the first 5-10 ml of blood, around 3 ml of blood were drawn using 4.3 ml citrate-plasma tubes and sent for analyses within 1 hour. The central laboratory of University Hospital Bern used the BIOPHEN^TM^ Heparin LRT Anti-Xa chromogenic assay and the BIOPHEN^TM^ Apixaban Calibrator, both HYPHEN BioMed®. Anti-Xa activity is determined via an accredited chromogenic substrate method and a proportional concentration output is provided in ng/mL via a serial dilution calibration process. Hereby, drug concentration is measured indirectly.

Patients were under apixaban 2.5 mg once or twice daily. Fixed time points of intake were assumed (morning = 08:00, midday = 14:00, night = 20:00). Time points of apixaban intake (morning, midday or evening) and start of dialysis sessions (morning = 08:00 or midday = 14:00) were not always synchronized. As a result, blood sampling before dialysis resulted in various intervals between intake and drug level monitoring. For those taking apixaban once daily (morning, midday or evening), drug level monitoring before dialysis (morning or midday) resulted in either 6, 12, 18 or 24 hours levels. For those taking apixaban twice daily (morning and evening), drug level monitoring before dialysis (morning or midday) resulted in either 6 or 12 hours levels. Whenever trough level obtainment was possible (once daily intake morning or midday with corresponding start/rhythm of dialysis; twice daily intake with morning not midday rhythm of dialysis), the dialysis care team routinely instructed patients in advance to postpone drug intake until blood had been sampled.

### Dialysis parameters

We obtained dialysis vintage at first drug level measurement, number and lengths of dialysis sessions, mode of dialysis (hemodialysis or hemodiafiltration, the latter being defined as a treatment with an exchange volume of ≥ 12 L per session), dialyzer specifications (membrane material, surface area), weight pre- / post dialysis and machine-based Kt/V.

### Calculation of Ultrafiltration-adjusted standard Kt/V

1. We took machine-calculated single pool Kt/V (spKt/V) from the week before drug level monitoring (3 sessions) and calculated mean weekly spKt/V.
2. We calculated equilibrated Kt/V (eKt/V) from spKt/V by using the equation suggested by Tattasall ^14^ with a slightly modified time constant (30.7 instead of 35 minutes) suggested by Daugirdas ^15^ based on results from the HEMO study.^16, 17^
3. We applied the Leypoldt equation ^18^ to calculate fixed-volume standard Kt/V (fv stdKt/V).
4. We adjusted fv stdKt/V for volume removal (ultrafiltration adjusted standard Kt/V; UF adj. stdKt/V) by using the Frequent Hemodialysis Network (FNH) equation ^19^ and estimated volume of urea distribution as 90% of the sex-adjusted Watson volume of total body water.

### Co-Variates

We included indication of apixaban, age, sex, BMI at first apixaban measurement, episodes of bleeding (via chart review) and antiplatelet therapy as co-variates.

### Statisical Analyses

We used STATA, version 17.0. We characterized population and results by mean/SD (age, BMI, dialysis dosage), median/IQR/minimum-maximum (drug level) or number/% (sex, dialysis modality, dialyzer brand, dialyzer surface). We calculated number of drug level measurements overall, per dosing group and per patient. We analyzed drug levels for the total population, for the two dosing groups and for the time points after intake (6, 12, 18 and 24 hours). For between group - comparisons we applied mixed-effects models to account for repeated measures within patients. We performed mixed effects multivariable linear regression analyses with drug level as dependent variable. For the independent variables we sequentially added anthropometrics, time-point of drug level measurement and dialysis parameters. Individual participants were included as random effect parameter. We log-transformed drug level values to achieve normal distribution of residuals and an improved fit of the model. For bleeding episodes we performed mixed effects multivariable linear regression analyses with drug level as dependent variable. Bleeding episodes according to concomitant anti-platelet therapy where evaluated using Fisher’s exact test. Due to the low number of participants and bleeding events, we refrained from investigating bleeding episodes in Cox or logistic models.

### Ethics

The study was approved on July 27, 2022 by the Cantonal Ethics Committee for Research of the Health, Social and Integration Directorate Bern, Switzerland. The project-ID is 2022-00981. At all stages, the study was executed according to the principles defined in the Declaration of Helsinki. All patients gave their informed consent to further use of health-related data.

## Results

### Characteristic of study population

We analyzed results from 24 hemodialysis patients. Mean (SD) age was 64.2 (15.3) years, 45.2 % were female and mean (SD) BMI was 27.9 (6.8) kg/m^2^. All patients dialyzed thrice weekly with a mean (SD) UF-adjusted standard Kt/V of 2.12 (0.26), using 5 different dialyzers and a wide range of dialyzer surface areas (Table 1).

**Table 1.**
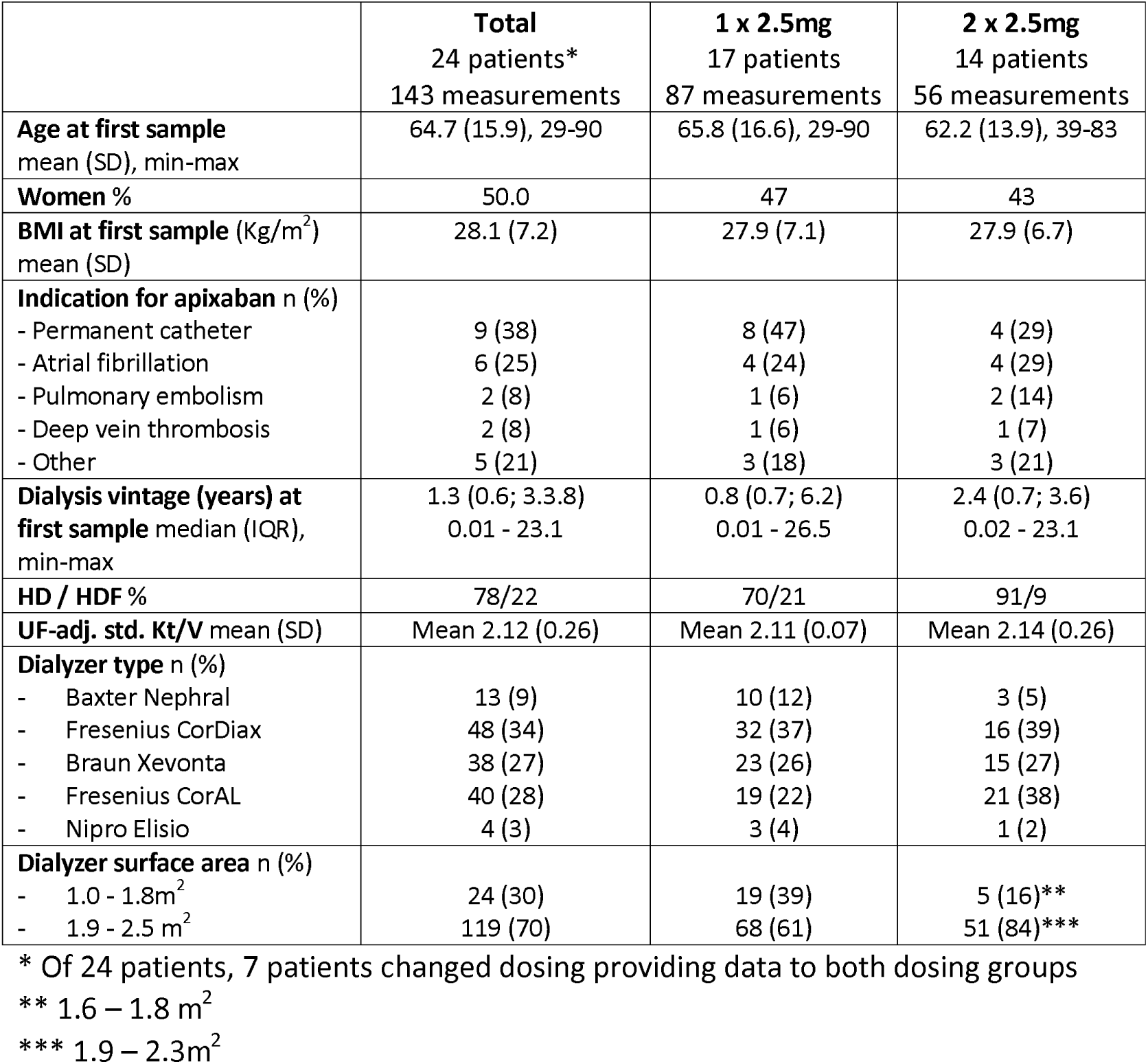
Characteristics of study participants, indication of apixaban and dialysis parameters.

|  | **Total**<br>24 patients*<br>143 measurements | **1 x 2.5mg**<br>17 patients<br>87 measurements | **2 x 2.5mg**<br>14 patients<br>56 measurements |
| --- | --- | --- | --- |
| **Age at first sample**<br>mean (SD), min-max | 64.7 (15.9), 29-90 | 65.8 (16.6), 29-90 | 62.2 (13.9), 39-83 |
| **Women %** | 50.0 | 47 | 43 |
| **BMI at first sample (Kg/m<sup>2</sup>)**<br>mean (SD) | 28.1 (7.2) | 27.9 (7.1) | 27.9 (6.7) |
| **Indication for apixaban n (%)** |  |  |  |
| - Permanent catheter | 9 (38) | 8 (47) | 4 (29) |
| - Atrial fibrillation | 6 (25) | 4 (24) | 4 (29) |
| - Pulmonary embolism | 2 (8) | 1 (6) | 2 (14) |
| - Deep vein thrombosis | 2 (8) | 1 (6) | 1 (7) |
| - Other | 5 (21) | 3 (18) | 3 (21) |
| **Dialysis vintage (years) at first sample** median (IQR), min-max | 1.3 (0.6; 3.3.8)<br>0.01 - 23.1 | 0.8 (0.7; 6.2)<br>0.01 - 26.5 | 2.4 (0.7; 3.6)<br>0.02 - 23.1 |
| **HD / HDF %** | 78/22 | 70/21 | 91/9 |
| **UF-adj. std. Kt/V** mean (SD) | Mean 2.12 (0.26) | Mean 2.11 (0.07) | Mean 2.14 (0.26) |
| **Dialyzer type n (%)** |  |  |  |
| - Baxter Nephral | 13 (9) | 10 (12) | 3 (5) |
| - Fresenius CorDiax | 48 (34) | 32 (37) | 16 (39) |
| - Braun Xevonta | 38 (27) | 23 (26) | 15 (27) |
| - Fresenius CorAL | 40 (28) | 19 (22) | 21 (38) |
| - Nipro Elisio | 4 (3) | 3 (4) | 1 (2) |
| **Dialyzer surface area n (%)** |  |  |  |
| - 1.0 - 1.8m<sup>2</sup> | 24 (30) | 19 (39) | 5 (16)** |
| - 1.9 - 2.5 m<sup>2</sup> | 119 (70) | 68 (61) | 51 (84)*** |

\* Of 24 patients, 7 patients changed dosing providing data to both dosing groups
\*\* 1.6 – 1.8 m<sup>2</sup>
\*\*\* 1.9 – 2.3m<sup>2</sup>

### Apixiaban drug level

143 drug levels were obtained during August 2017 and January 2023. From 24 patients, 7 patients changed dosing during observation period providing data to both dosing regimens. 17 patients were on 2.5 mg once daily providing 87 levels with a mean (SD) of 4.9 (4.1) measurements per case. 14 patients were on 2.5 mg twice daily providing 56 levels with a mean (SD) of 3.8 (2.8) measurements per case (Table 2). Median (IQR) follow-up defined as time between first and last drug level measurement was 12.5 (5.5 - 21) months with a maximum follow-up of 51 months.

**Table 2.** Apixaban drug level for the total population and the two dosing groups.

|  | **Total** | **1 x 2.5mg** | **2 x 2.5mg** |
| --- | --- | --- | --- |
| **No of patients n** | 24 * | 17 | 14 |
| **No of samples n** | 143 | 87 | 56 |
| **No of samples per case mean (SD), min-max** | 4.5 (3.7), 1-17 | 4.9 (4.1) 1-17 | 3.8 (2.8), 1-11 |
| **% of samples pre/post dialysis** | 92/8 | 91/9 | 93/7 |
| **Drug level, ng/ml(all)** |  |  |  |
| - Median (IQR) | 56.0 (40.9 - 82.4) | 54.1 (< 40 - 72.1) | 71.3 (45.8 - 104.1) |
| - 5/95 percentile | < 40/127.1 | < 40/118.7 | < 40/140.5 |
| - Min. - Max. | <40 - 223.7 | <40 - 157.6 | < 40 - 223.7 |
| - % Below detection limit | 26.0 | 29.9 | 14.3 |
| **Drug level, ng/ml, (trough) n** | 85 | 52 | 33 |
| - Median (IQR) | 54.1 (40.0 - 78.6) | 51.8 (< 40 - 70.0) | 70.4 (45.5 - 102.7) |
| - 5/95 percentile | < 40/133.4 | < 40/114.8 | < 40/173.4 |
| - Min. - Max. | <40 - 223.7 | < 40 - 127.1 | < 40 - 223.7 |
| - % Below detection limit | 27.1 | 36.5 | 12.1 |

\* Total number of patients is 24. Total number of cases is 31 as 7 patients are in both dosing group as dosing was changed during treatment period. Drug level (all) of twice daily dosing are significantly higher than those of the once daily dosing. Trough level and non-through level within each dosing group were not significantly different from each other.

Median (IQR) apixaban drug level were 54.1 (< 40 - 72.1) ng/mL in the 2.5 mg once daily and 71.3 (45.8 - 104.1) ng/mL in the twice daily group, P < 0.001 (Table 2, Figure 1a). Maximum level were 157.6 ng/mL and 223.7 ng/mL respectively. 30 % of drug level in the once daily and 14 % in the twice daily dosing group were below our assays detection limit of < 40 ng/mL. Results are presented in Table 2. Drug level pre and post hemodialysis did not differ from each other. For both dosing groups, trough level (taken 24 hours after the once daily and 12 hours after the twice daily dosing) compared to non-trough level (taking 6, 12 or 18 hours after the once daily and 6 hours after the twice daily dosing) were not significantly different from each other (Figure 1b). Also, drug level measurements at different time points did not differ significantly from one another (Figure 1c and 1d).

**Figure 1.**
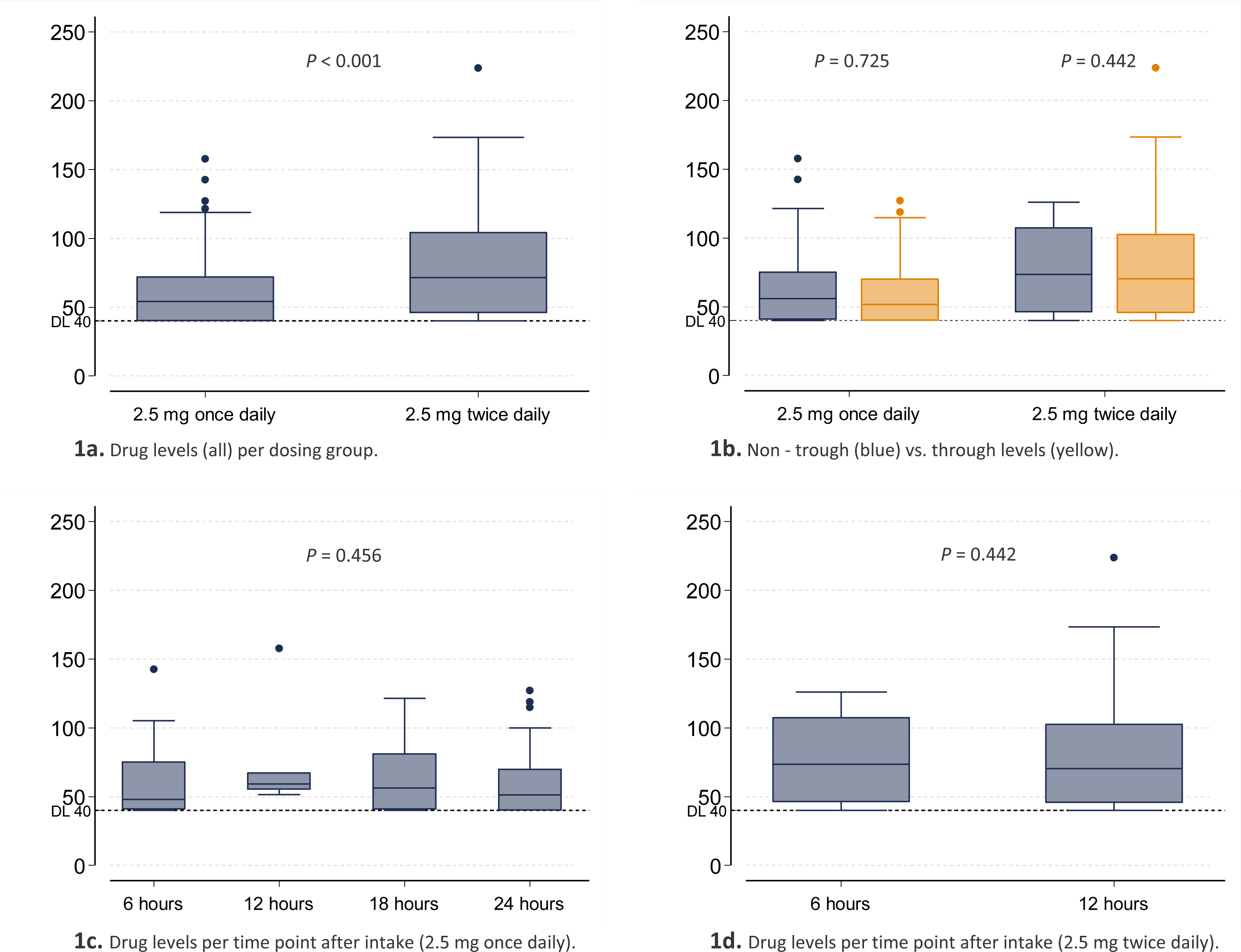
Apixaban drug level. Comparisons of dosing groups, non - trough versus trough level and time points of intake. DL, Detection Limit.

### Mixed-effects models analysis

In the mixed-effects multivariable linear regression analysis, only dosing group (2.5 mg twice daily higher than once daily) was significantly associated with log apixaban drug level (Table 3). Sensitivity - Analyses including dialyzers specificity (Acrylonitrile versus PES plus PSU or Acrylonitrile versus PES versus PSU) or adding dialyzer surface as a dichotomous variable (1- 1.8 versus 1.9 - 2.5 m^2^) did not change results.

**Table 3.**
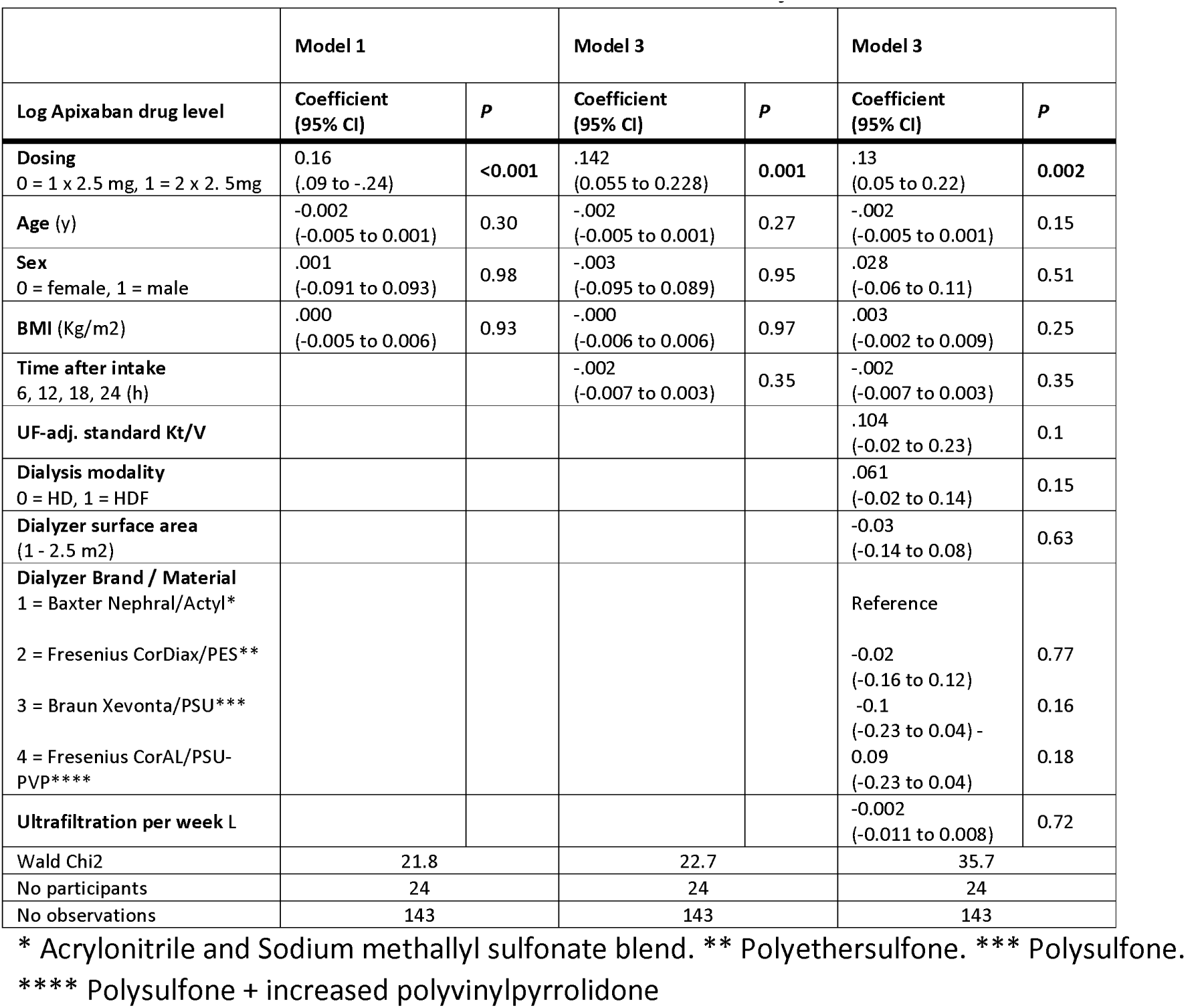
Mixed-effects model with 3 level of multivariate adjustments.

|  | Model 1 |  | Model 3 |  | Model 3 |  |
| --- | --- | --- | --- | --- | --- | --- |
| Log Apixaban drug level | Coefficient<br>(95% CI) | P | Coefficient<br>(95% CI) | P | Coefficient<br>(95% CI) | P |
| Dosing<br>0 = 1 x 2.5 mg, 1 = 2 x 2.5 mg | 0.16<br>(.09 to -.24) | <0.001 | .142<br>(0.055 to 0.228) | 0.001 | .13<br>(0.05 to 0.22) | 0.002 |
| Age (y) | -.002<br>(-0.005 to 0.001) | 0.30 | -.002<br>(-0.005 to 0.001) | 0.27 | -.002<br>(-0.005 to 0.001) | 0.15 |
| Sex<br>0 = female, 1 = male | .001<br>(-0.091 to 0.093) | 0.98 | -.003<br>(-0.095 to 0.089) | 0.95 | .028<br>(-0.06 to 0.11) | 0.51 |
| BMI (Kg/m2) | .000<br>(-0.005 to 0.006) | 0.93 | -.000<br>(-0.006 to 0.006) | 0.97 | .003<br>(-0.002 to 0.009) | 0.25 |
| Time after intake<br>6, 12, 18, 24 (h) |  |  | -.002<br>(-0.007 to 0.003) | 0.35 | -.002<br>(-0.007 to 0.003) | 0.35 |
| UF-adj. standard Kt/V |  |  |  |  | .104<br>(-0.02 to 0.23) | 0.1 |
| Dialysis modality<br>0 = HD, 1 = HDF |  |  |  |  | .061<br>(-0.02 to 0.14) | 0.15 |
| Dialyzer surface area<br>(1 - 2.5 m2) |  |  |  |  | -0.03<br>(-0.14 to 0.08) | 0.63 |
| Dialyzer Brand / Material<br>1 = Baxter Nephral/Actyl*<br>2 = Fresenius CorDiax/PES**<br>3 = Braun Xevonta/PSU***<br>4 = Fresenius CorAL/PSU-pVP**** |  |  |  |  | Reference<br>-0.02<br>(-0.16 to 0.12)<br>-0.1<br>(-0.23 to 0.04) -<br>0.09<br>(-0.23 to 0.04) | 0.77<br>0.16<br>0.18 |
| Ultrafiltration per week L |  |  |  |  | -0.002<br>(-0.011 to 0.008) | 0.72 |
| Wald Chi2 | 21.8 |  | 22.7 |  | 35.7 |  |
| No participants | 24 |  | 24 |  | 24 |  |
| No observations | 143 |  | 143 |  | 143 |  |

\* Acrylonitrile and Sodium methallyl sulfonate blend. \*\* Polyethersulfone. \*\*\* Polysulfone.
\*\*\*\* Polysulfone + increased polyvinylpyrrolidone

### Bleeding events

In 7 out of 24 patients (29%) a bleeding event was documented with 8 bleeding events in total (one patient had two episodes): Masseter bleeding (apixaban 2.5 mg once daily, plus antiplatelet); splenic bleeding with hemorrhagic shock (apixaban 2.5 mg once daily, plus antiplatelet, apixaban stopped); upper gastrointestinal bleeding - circulatory stable (apixaban 2.5 mg once daily, plus antiplatelet and NSAID, antiplatelet stopped); hemorrhagic pericardial effusion with hemodynamic impairment (apixaban 2.5 mg once daily); lower gastrointestinal bleeding - circulatory stable (apixaban 2.5 mg twice daily, plus antiplatelet); two mild bleedings from prostate varicose veins (apixaban 2.5 mg twice daily, plus antiplatelet, apixaban dose reduction, later stopped, bleeding recurred without apixaban but antiplatelet therapy only); mild bleedings from rectal condylomata (apixaban 2.5 mg twice daily, plus antiplatelet, apixaban dose reduction).

Mean apixaban levels of patients with versus without a bleeding event where not significantly different: 76.3 (SD 24.3) versus 61.7 (SD 23); P = 0.18. However, apixaban drug levels measured before the bleeding event were significantly higher compared to drug levels without a subsequent bleeding event: 115 (SD 51.6) versus 65.9 (SD 31.6) ng/mL; difference 51.2 ng/mL (95%-CI 31.4 to 71.1); P < 0.001.

From 7 patients with a bleeding episode, 6 patients (86%) took concomitant antiplatelet therapy whereas from 17 patients without bleeding events only one patient (6%) was under additional antiplatelet therapy (P < 0.001).

### Change of apixaban dosing due to drug level

In 6 out of 24 patients (25%) the treating physicians decided to change apixaban dosing regimen due to drug level results. In three patients, 2.5 mg once daily dosing was increased to twice daily dosing, with resulting mean (SD) drug level changes from 46.2 (6.2) to 52.2 (1.6), < 40 to 96.5 (19.8) and 43.4 (5.1) to 101.3 ng/mL. In two patients, 2.5 mg twice daily dosing was reduced to once daily dosing, with a resulting mean (SD) drug level change from 78.6 (28.6) to 64.8 (4.5) and 134.1 (20.3) to 56.4 (8.2) ng/mL. In one further patient, a trial of twice daily dosing resulted in a drug level of 173 ng/mL with drug level for the once daily dosing regimen before and thereafter of 83.5 (15.3) and 90.8 (15.9) ng/mL respectively.

### Change of apixaban dosing due to bleeding

After 3 out of the 8 bleeding episodes, apixaban medication was adjusted. In one episode, reduction from 2.5 mg twice to once daily dosing resulted in a mean (SD) drug level change from 119.7 (1.1) to 52.7 (4.1) ng/mL. After 2 bleeding episodes apixaban was stopped.

### Drug level over time

On an individual patient level, we did not see signs of accumulation in 25 cases that provided at least 2 measurements (Figure 2). In these patients, first drug level measurement took place at a median (IQR) of 14 (8 - 30) days after initiation of apixaban (available data for 15 cases) with a total follow-up time of median (IQR) 12 (2 - 16) and a range of 1 - 51 months intake (available data for all cases).

**Figure 2.**
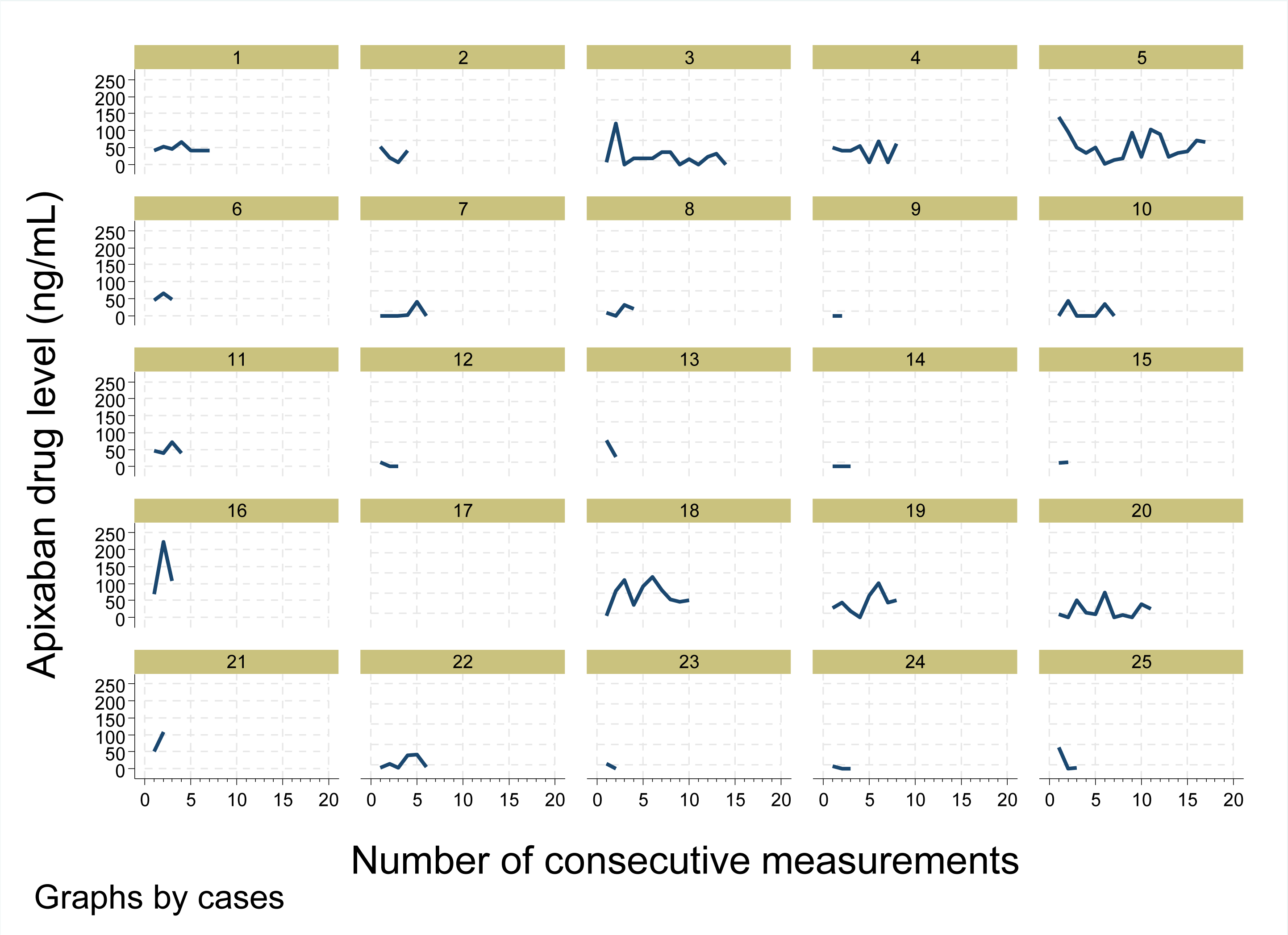
Apixaban level (all) of each participant over time. Serial drug level monitoring from 25 hemodialysis patients each providing between 2 and 17 drug levels. First drug level measurement took place at a median (IQR) of 14 (8 - 30) days after initiation of apixaban (available data for 15 cases) with a total follow-up time of 12 (2 - 16) months intake (available data for all cases).

### Drug level in comparison

In Figure 3 and Table 4 scatter plots of our drug level are compared to 10th to 90th and 5th to 95th percentiles of drug level from non-CKD patients taking 5 mg apixaban twice daily ^20–22^.

**Figure 3.**
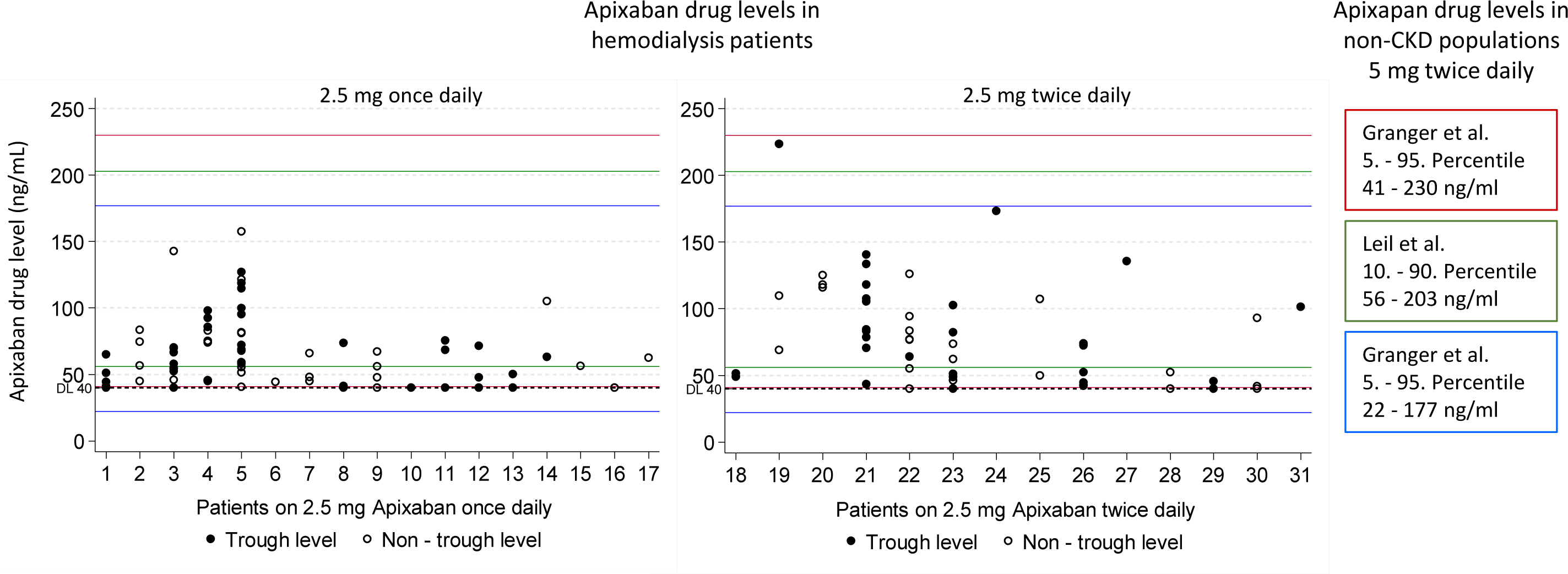
Comparison of measured apixaban levels in our study with those from non-CKD patients taking therapeutic doses of 5mg twice daily. Cases 1 - 17 were on 2.5 mg once daily, cases 18-31 were on 2.5 mg apixaban twice daily. Apixaban levels were measured on dialysis days. From 143 samples, 85 were trough level and 58 were non-through level. Trough and non- through level were not significantly different from each other. The lower detection limit of our assay was 40 ng/mL. Comparative confidence intervals (Boxes on the right side as well as reference lines in red, green and blue) are derived from apixaban trough levels non-CKD populations ^20–22^ provided by the European Union summary of product characteristics.^8^

**Table 4.** Apixaban level in comparison to the literature in HD and non-CKD populations

|  | Hemodialysis |  | Non-CKD |  |  |
| --- | --- | --- | --- | --- | --- |
| Author Year<br>(no. patients), prospective<br>pharmacokinetic study = P | Apixaban level (ng/ml) |  | Apixaban level (ng/ml) |  | Author Year<br>Observed (o), predicted (p) |
|  | Median<br>(<sup>1</sup>mean) | 5/10<sup>th</sup> percentile<br>(<sup>2</sup>range, <sup>3</sup>2 SD) | Median | 5/10<sup>th</sup> percentile<br>(<sup>2</sup>range, <sup>4</sup>10/90<sup>th</sup>) |  |
|  | 2.5 mg once daily |  | 2.5 mg once daily |  |  |
| Our data 2023 (14) | 54 | <40/119 |  |  |  |
|  | 2.5 mg twice daily |  | 2.5 mg twice daily |  |  |
| Roberge 2017 (1) | NA | 58/84 | NA | 16/27 | Yamahira 2014 (o) |
| Mavrakanas 2021 (7), P | 58<sup>1</sup> | 22/94<sup>3</sup> | 56 | 24/103<sup>4</sup> | Leil 2010 phase II (o) |
| Our data 2023 (17) | 71 | <40/141 | 79 | 34/162 | Cirincione 2018 phase I-III (p) |
|  |  |  | 63 | 11/90 | Byon 2017 (p) |
|  |  |  | 85 | 41/200<sup>2</sup> | Mavri 2021 (o) |
|  | 5 mg twice daily |  | 5 mg twice daily |  |  |
| Mavrakanas 2017 (5), P | 218 | 91/337<sup>2</sup> | NA | 49/78 | Yamahira 2014 (o) |
| Pokorney 2022 (42), P*, ** | 142 | NA | 107 | 56/203<sup>2</sup> | Leil 2010 Phase II (o) |
| Pokorney 2022 (21), P*, *** | 168 | NA | 103 | 41/230 | Cirincione 2018 phase I-III (p) |
|  |  |  | 120 | 22/177 | Byon 2017 (p) |
|  |  |  | 117 | 63/291<sup>2</sup> | Mavri 2021 (o) |
|  | 10 mg twice daily |  | 10 mg twice daily |  |  |
|  |  |  | NA | 104/114 | Yamahira 2014 (o) |
|  |  |  | 120 | 41/335 | Byon 2017 (p) |

\* The cohort with pharmacokinetic data comprised in total 63 patients. 43 patients took 5 mg twice daily. 20 patients took 2.5 mg twice daily of which 14 met dose reduction criteria of age ≥80 years and/or weight ≤60 Kg. \*\* Group (42 patients) without major or clinically relevant non-major bleeding. \*\*\* Group (21 patients) with major or clinically relevant non-major bleeding. P = prospective pharmacokinetic study. o = observed. p = predicted.

### Race and Ethnicity

In this study, one participant (4%) self-identified as Hispanic, 23 participants (96%) self-identified as White.

## Discussion

We provide results from apixaban drug level monitoring in hemodialysis patients on different dosing regimens. We investigate risk factors for increased drug level, an analysis with regard to bleeding events and we provide a literature comparison regarding non-CKD populations.

The FDA recommendation for apixaban dosing is 5 mg twice daily for both, non-CKD and hemodialysis patients.^9^ However, prospective pharmacokinetic data in hemodialysis suggest that 2.5 mg twice daily result in similar drug level like non - CKD populations under 5 mg twice exceeding their 95^th^ percentile if 5 mg twice daily is applied.^23^ Regarding safety and efficacy, randomized controlled trial (RCT) data showed no differences to vitamin k antagonists with 2.5 mg twice daily in hemodialysis patients.^10^ However, even with 2.5 mg twice daily, serious bleeding complications have been reported ^12^.

With 2.5 mg twice daily, our median drug levels (71 ng/mL) were lower than those of non-CKD populations taking 5 mg twice daily (103, 107, 120 ng/mL)^20–22^, published among others by the European Union summary of product characteristics.^8^ Our 95^th^ percentiles and even our three highest single drug levels (224, 173, 157 ng/mL) did not exceed those of non-CKD reference populations. Hence, looking at drug level data only, our results suggest, that 2.5 mg twice daily is not associated with overtly increased drug exposition compared to non-CKD patients for whom licensure studies were performed.

On the contrary, our results rather suggest, that with 2.5 mg twice daily, and even more with 2.5 mg once daily dosing, individual patients might be under-dosed regarding sufficiently protective drug exposure, arguing for more routine drug monitoring. It is worth mentioning that 2.5 mg once daily dosing might still confer valuable prophylactic benefits with regard to long-term patency of venous access for dialysis treatment.

Comparing drug levels of our 2.5 mg twice daily group with non-CKD populations taking the same 2.5 mg twice daily dose,^20–22^ results were roughly comparable (median: 71 ng/mL versus 56, 79, 63 ng/mL; 5^th^ percentile: < 40 ng/mL versus 24, 34, 11 ng/mL; 95^th^ percentile: 141 ng/mL versus 103, 162, 90 ng/mL).

Pokorney et al. found that AUC0-12 from 5 mg twice daily dosing in hemodialysis patients was significantly higher compared to non-CKD participants of the ARISTOTLE trial (2475 ng/mL×h versus 1374 ng/mL×h) but comparable to ARISTOTLE participants with an eGFR of 15 - 59 ml/min/1.73m^2^.^24^ These results suggest that in hemodialysis, 5 mg twice daily dosing can be both, appropriate or too high, arguing for contributory drug monitoring.

Trough levels did not differ from non-trough levels. Important factors here are the retrospective nature of the study and the considerable intra- and inter-individual variabilities, which are well-known challenges of apixaban drug levels monitoring.^1, 25^ However, even under these real-live conditions, no exacerbation of drug levels were seen.

Existing literature does not support valid assessment of efficacy or bleeding risk via drug monitoring ^24–26^ and our analysis of increased bleeding events with higher drug levels or concomitant antiplatelet therapy is clearly exploratory. However, as long as valid outcome data are scarce, and as there is data on the adequacy of both, twice daily 2.5 mg or 5 mg dosing,^3, 9, 10, 24^ our findings are a contribution with regard to drug monitoring as an additional mean to increase safety.

Mixed-effects models multivariable analyses, did not show age, sex, BMI or time point of measurement after drug intake to be independently associated with drug levels. Prospective pharmacokinetic studies in hemodialysis patients considering age, sex and BMI differences are missing. Regarding non-hemodialysis populations, pharmacokinetic studies have shown the following: Older patient showed similar or slightly higher level than young patients.^27^ However, age did not appear to influence exposure-response relationship,^21^ and thus did not justify a dose reduction as a single parameter.^9^ Frost et al. found 18% and 15% higher Cmax and AUC values in healthy female compared to male patients which was considered a modest difference, unlikely to be clinically meaningful.^27^ Higher drug level values (AUC +20%) were found in low body weight (< 50 Kg) and lower values (AUC -23%) in high body weight (>120 Kg).^28^ Lower bodyweight is incorporated not as a single but one out of 3 dose reduction factors recommended by the FDA.^9^

Drug levels at different time points after intake were not significantly different. This result has to be interpreted with caution, as values represent the whole study population but not a pharmacokinetic analysis of single patients. None of the dialysis-associated parameter were independently associated with drug levels. This was somewhat unexpected. Apixaban has a molecule size of 495.5 Dalton, a protein binding of 87% and a distributional volume of 21 L.^9^ Thus, a fractional removal by hemodialysis is expected. Small prospective pharmacokinetic studies show a reduction in AUC by hemodialysis of -14% (pre/post) ^3^ and -26% (AUC-48h) ^4^ for a 5 mg dose and a AUC-48h reduction of -48% for a 2.5 mg dose.^4^ Potentially, our inter-individual differences in dialysis dose were too small. However, in light of low sample numbers and retrospective analyses, interpretation of multivariable regression results is limited. Intake of a second dose of 2.5 mg of apixaban per day was consistently associated with higher drug level. This highlights at least a rough relationship between amount of intake and steady state drug level supporting drug monitoring as a potential additional mean for dose finding on an individual basis.

Our analysis does not suggest drug accumulation over time independent of dosing regimen and length of follow-up. In contrast, pharmacokinetic analyses revealed significant accumulation in hemodialysis patients during the first 8 days.^23^ Our data do not allow valid analyses of drug level within the first few days after starting the drug. However, with regard to our follow-up period (IQR 5.5 - 21 months), our data clearly support the picture of a non-accumulating steady state.

Our study has several strengths. Compared to the existing literature, we provide a high number of drug levels. We integrate different dosing regimens and provide follow-up data of more than 5 years. We incorporate detailed dialysis-related co-variates in our analyses. We include an analysis of bleeding events and we provide a comparative review to drug levels from non-CKD populations facilitating interpretation of drug levels and supporting sensible decision making regarding appropriate dosing.

Our study has several limitations. The analysis of bleeding events is exploratory. Furthermore, retrospective nature of the study limits precise specification of timing of drug intake. Hence, results from multivariable analyses need to be interpreted with caution. Drug levels in general, however, do picture results from a tertiary care real-life setting.

In conclusion our analyses suggest, that drug level monitoring can be a contributory tool to increase safety and appropriate dosing in hemodialysis patients on apixaban. As results from drug monitoring have not been proven to reliably picture drug efficacy or bleeding risk, this primarily applies to dosing adjustments in case of overtly low/non-detectable or overtly high dug levels compared to confidence intervals of non - CKD reference populations. Drug levels from hemodialysis patients taking 2.5 mg twice daily did not exceed those from non - CKD populations taking 5 mg twice daily. There might be individual cases of relevant under-dosing with 2.5 mg once daily and, to a lesser extent, also with 2.5 mg twice daily dosing. Higher drug levels and concomitant antiplatelet therapy might result in higher risks of bleeding. Further prospective pharmacokinetic evidence of larger scale is needed to clarify if valid reference values for drug level can be found translating to dosing recommendation regarding a beneficial balance of sufficient anticoagulation and likelihood of bleeding.

## Disclosure

Financial conflict of interest: Simeon Schietzel: none, Andreas Limacher: none, Matthias B. Moor: none, Cecilia Czerlau: none, Bruno Vogt none, Fabienne Aregger: none, Dominik E. Uehlinger: none.

## Authors‘ Contributions

Simeon Schietzel: Conceptualization, methodology, funding acquisition, formal analysis, visualization, writing of manuscript. Andreas Limacher: Methodology, formal analysis, validation, review of manuscript. Matthias B. Moor: Supervision, review of manuscript. Cecilia Czerlau: Conceptualization, review of manuscript. Bruno Vogt: Supervision, review of manuscript. Fabienne Aregger: Supervision, review of manuscript. Dominik E. Uehlinger: Funding acquisition, Supervision, review of manuscript.

## Data Availability

Raw data will be made available upon reasonable request.

## Data Availability

Data will be made available upon reasonable request to the corresponding author.

## Acknowledgments

Special thanks to the dialysis care team of the department of nephrology and hypertension of the University Hospital of Bern for their invaluable contribution to the apixaban drug level monitoring program.

